# SARS-CoV-2 Antibody Response during Omicron Predominance after COVID-19 Vaccination in People Living with HIV: A Comparative Study in Canada and Burkina Faso

**DOI:** 10.64898/2026.05.26.26354060

**Authors:** Hend Jarras, Wilfried W. Bazié, Isalie Blais, Arielle Pakenham, Justin Valiquette, Mathieu Thériault, Isidore T. Traore, Dramane Kania, Aline Raissa Ouoba, Ivette Zoundi, Martin Pelletier, Philippe A. Tessier, Marc Pouliot, Sylvie Trottier, Marie-Louise Vachon, Caroline Gilbert

## Abstract

People living with HIV (PLWH) are known to maintain a degree of immune deficiency despite efficient antiretroviral therapy and may exhibit diminished responses to vaccines. In this study, we assessed the immune response to SARS-CoV-2 infection and vaccines in two geographically distinct PLWH populations. PLWH and HIV-negative (HIV-) participants were recruited from Quebec City (QC), Canada, and Bobo-Dioulasso (BD), Burkina Faso, for two visits at 24-week intervals during the predominance of the Omicron variant, from May 2022 to September 2023. Blood samples were collected at each visit for the detection of antibodies against spike (anti-S) and nucleocapsid (anti-N) proteins of SARS-CoV-2 in platelet-free plasma. A total of 360 participants were enrolled. We detected anti-S antibodies in 99% of participants, indicating that nearly all had prior exposure to the SARS-CoV-2 spike antigen, either through vaccination or prior infection. Anti-S titers showed no difference between PLWH and HIV-participants in each location, while significantly higher titers were observed in participants from QC compared to BD. In contrast, anti-N antibodies, indicative of prior infection, were detected in 39% and 86% of the participants in QC and BD, respectively, suggesting that the virus circulated largely in the latter population. No difference in anti-N levels was observed between PLWH and HIV-participants in BD. However, participants in QC had significantly lower titers compared to HIV-participants. Overall, this study shows that PLWH develop robust antibody responses to SARS-CoV-2 vaccination, comparable to those observed in HIV-participants. Significant geographic differences were observed in anti-S titers, irrespective of HIV status, with participants from QC displaying higher titers. In contrast, participants from BD had higher anti-N antibody prevalence and titers, reflecting more SARS-CoV-2 infections in BD than in QC. Finally, analysis of anti-S antibody titers against several circulating variants revealed significantly lower levels in unvaccinated participants and in those vaccinated with monovalent vaccines in BD. No significant difference was observed between monovalent and bivalent vaccines administered in QC.

## INTRODUCTION

The Coronavirus Disease 2019 (COVID-19) pandemic has challenged healthcare systems, economies, and community well-being worldwide (1). In countries with well-established public health infrastructures, coordinated epidemiological efforts have enabled large-scale screening and efficient case identification. In sub-Saharan African countries, limited access to screening, fragile health systems, and the stigmatization of confirmed and contact cases have hindered diagnosis and contributed to the perception that these populations were, to some extent, less susceptible to infection (2–4). While protection against symptomatic and severe forms of the disease has been confirmed in sub-Saharan African countries, sero-surveillance data indicate that the virus has circulated widely in these populations, with prevalence exceeding 50% in some countries by the end of 2021 (3, 5, 6).

The approval and deployment of vaccines changed the pandemic landscape, particularly regarding the incidence of severe disease (7, 8). Initially developed as a single-dose vaccine, the World Health Organization (WHO) later recommended additional doses to boost the immune response following the emergence of new variants, such as the Alpha and Delta variants. However, some of these variants increasingly showed resistance to neutralization by antibodies generated through prior vaccination or infection, prompting manufacturers to adapt vaccines accordingly (9, 10). A few months after the emergence of the Omicron variant, different subvariants of the original mutant emerged, broadening the diversity of circulating variants (11). Many were detected in Canada and were considered variants of concern, including subvariants BA.2, BA.4, BA.5, BQ.1, XBB, and JN.1 (12, 13). Surveillance was less extensive in sub-Saharan Africa, where few information on Omicron circulating subvariants is available (13). The BA.4 and BA.5 subvariants were more contagious than their predecessors (14), while the BQ and XBB sublineages were found to have an even greater capacity for immune evasion than the original Omicron variant (15). This led to the production of bivalent vaccines containing the BA.1 and BA.4/BA.5 Omicron subvariants in 2022 to respond to the increased transmissibility of those variants (16, 17).

The hastened pace of vaccine development meant that specific populations, including immunocompromised individuals and those with chronic diseases, were often underrepresented or excluded from clinical trials (18, 19). This created a lack of data on the immunogenicity and efficacy of SARS-CoV-2 vaccines in these populations even if early pandemic data indicated that immunocompromised individuals or those with chronic inflammatory diseases were at increased risk of complications from SARS-CoV-2 infection (20–24).

On a global scale, people living with HIV (PLWH) represent a diverse population with compromised immunity marked by immune activation and chronic inflammation (25, 26). Despite effective antiretroviral therapy (ART), PLWH show several innate and adaptive immune deficiencies (27). In fact, lower seroconversion rates have been reported in these patients, and antigen-specific antibody responses also present shorter half-life after immunization (28, 29). Persistent immune activation appears to contribute to both reduced antibody levels and impaired memory B cell responses following influenza vaccination (30). Studies on hepatitis A and B vaccination have shown that markers of advanced HIV disease, specifically CD4 counts below 200 cells/µL and a low CD4/CD8 ratio, correlate with weaker and less durable vaccine-induced immunity (31, 32). In a study on pneumococcal vaccination, low overall B-cell counts and memory B-cell depletion were associated with poor vaccine responses (33). Consequently, elucidating the magnitude and durability of the antibody response elicited by SARS-CoV-2 vaccination in PLWH is essential to optimize vaccination strategies for this population.

In this context, we aimed to assess PLWH immune response to SARS-CoV-2 vaccines and infection. We hypothesized that due to immune impairment, PLWH would develop weaker antibody responses following vaccination or infection, even in the presence of effective ART. To verify this hypothesis, we compared serological responses among PLWH with those of the general population in an observational study involving PLWH and HIV-negative (HIV-) controls from North America and sub-Saharan Africa. This article presents the study’s experimental design along with clinical, demographic, and serological findings.

## METHODS

### Design and Setting of the Study

In this observational study, antibody levels against SARS-CoV-2 spike and nucleocapsid antigens were measured at two visits spaced by 24 ± 2 weeks, in groups of PLWH and HIV-participants. The study took place in two different locations: Quebec City (QC), Canada, and Bobo-Dioulasso (BD), Burkina Faso. QC population had a high vaccination rate and well-documented SARS-CoV-2 infection and vaccine exposure, whereas BD is characterized by a lower vaccination rate and limited data on the extent of the SARS-CoV-2 infection.

### Participants

In QC, PLWH were recruited at the *“Unité hospitalière de recherche, d’enseignement et de soins sur le SIDA”* (UHRESS) of the *“CHU de Québec-Université Laval-site CHUL”*. HIV-volunteers were recruited from participants of an observational study on the seroprevalence of SARS-Co-2 infection (*Immunité cellulaire et séroprévalence des anticorps contre SARS-CoV-2: Caractérisation de trois populations de travailleurs de l’alimentation,* registration number CER-2021-5744) and who agreed to be part of a voluntary registry (*Registre de participants de recherche clinique en infectiologie du CHU de Québec-Université Laval,* registration number CER-2019-4163) (34–36). These HIV-participants were matched for age and sex with PLWH from QC. They were tested for HIV to confirm a seronegative condition. In BD, PLWH were recruited at Centre Muraz *Yerelon* Clinic and from other collaborating HIV care centers. The *Yerelon* Clinic also offers HIV screening services, through which HIV-participants were recruited. The study was approved by the ethics committee of the “*Centre de recherche du CHU de Québec-Université Laval”* (registration number CER-2022-6241) and the Burkina Faso Ministry of Health Research Ethics Committee (Deliberation n°2022-03-048). Each participant provided written informed consent before enrollment and was assigned an anonymous ID used for data and sample management. The inclusion criteria were as follows: 1) age 18 years or older; 2) ability to provide informed consent; 3) ability to travel to the recruiting centers; 4) residence in one of the study areas (QC or BD); and 5) confirmed HIV-1 status-positive for the PLWH group and negative for the control group. Exclusion criteria for all groups were as follows: 1) a positive SARS-CoV-2 detection test (PCR or Rapid Antigen) within the past 10 days; 2) contact with a person with a positive SARS-CoV-2 detection test in the last 14 days; 3) receiving any vaccine within the past 14 days; 4) anemia, defined as hemoglobin < 120 g/L in men, and < 100 g/L in women; 5) blood donation within the past four weeks; 6) pregnancy; and 7) any condition that, in the physician’s judgement, could interfere with study participation, lead to early withdrawal, or warrant discontinuation.

### Data Collection and Follow-up

At the first visit (V1), informed consent was obtained from eligible participants. A separate consent was obtained for the biobanking of samples and data for future research. All participants were then interviewed by nurses to collect data, including demographic information, medical history, current medication, HIV-1 serology, most recent HIV viral load, co-infections, and co-morbidities. Data were also collected on SARS-CoV-2 exposure, infection, diagnosis, and vaccination history. Participants then provided a venous blood sample. At the second visit (V2), participants completed a short questionnaire to update health events since the last visit and provided a venous blood sample.

### Blood Collection and Biobanking

The complete protocol for blood collection and processing is provided in **Appendix I** (Supplementary Material). Briefly, blood (120 mL) was drawn under aseptic conditions by nurses. Different types of tubes were used: 1) EDTA tubes for plasma, peripheral blood mononuclear cell (PBMC) and polymorphonuclear neutrophil (PMN) purification, and dried blood spots (DBS) confection; 2) dry tubes for serum; 3) citrate tubes for citrated plasma and PBMCs; and 4) heparin tubes for heparinized plasma and PBMCs. Platelet-free plasma, serum, frozen cells and activated cell supernatants were stored in the biobank (*“Biobanque pour la conservation de matériels biologiques et des données en lien avec l’étude de l’immunocompétence des personnes vivant avec le VIH-1”* registration number CER-2023-6445).

### Measurement of HIV-1 RNA

HIV-1 viral load was measured using the cobas^®^ 8800 System (Roche Molecular Systems, Inc., New Jersey, USA) on plasma samples with a limit of detection of 13.2 copies/mL and a range of quantification of 20 copies/mL to 1 x 10^7^ copies/mL in QC. In BD, HIV-1 viral load was measured using the Cobas AmpliPrep /Cobas TaqMan real-time PCR Assay (TaqMan, Roche Diagnostics, Mannheim, Germany), with a detection limit of 20 copies/mL.

### CD4^+^ and CD8^+^ T Lymphocyte Counts

Absolute counts of CD4^+^ and CD8^+^ T lymphocytes were obtained using Multitest CD45/CD3/CD8/CD4 kits on a FACSCanto II (BD Biosciences, San Diego, CA) in QC. A single-platform approach using BD Biosciences Trucount absolute-count tubes was employed. In BD, absolute counts of CD4^+^ and CD8^+^ T lymphocytes were obtained using a Partec CyFlow Counter system (Sysmex Partec GmbH, Arndtstr. 11 a-b, 02826 Görlitz, Germany), with CD4 Easy Count Kit and CyFlow CD8 PE-Cy5 antibodies. The complete blood count was performed using a Sysmex XN-330^TM^ automated hematology analyzer (Sysmex Corporation, Kobe, Japan).

### SARS-CoV-2 Antibody Assessment

The test was first run with 12 pre-COVID and pre-Omicron plasma samples to establish positivity thresholds for all antigens. Plasma samples were obtained from 2 biobanks: the “*Cellules et nanovésicules sanguines des patients VIH-1 positifs et non VIH-1*” (CER-12-03-167) and; (CER-2021-5744). The average antibody titer was calculated, and the threshold for a positive result was set at 3 standard deviations above the mean. Stored citrated platelet-free plasma samples were thawed at 4°C and incubated overnight with a 1 mM 2,2’-dithiodipyridine (AT-2) (Sigma, Cat. D5767-5G) solution to inactivate HIV-1 (37). HIV-samples were also treated to ensure comparability between groups. Antibodies against the SARS-CoV-2 nucleocapsid and 9 spike variants (ancestral, BA.1, BA.5, BQ.1.1, XBB.1, XBB.1.16, XBB.1.16.1, XBB.1.5, XBB.2.3) were measured using Panel 36 of the V-PLEX COVID-19 Serology kit from Meso Scale Discovery (Cat. K15715U-2). The manufacturer’s recommended protocol was followed. Briefly, samples were diluted 1:20,000 in Diluent 100. Plates were incubated with Blocker A solution for 30 min at room temperature (RT) with shaking. After washing, samples, standards, and controls were added to the plates and incubated at RT with shaking for 2h. The plates were washed and incubated in the dark with the detection antibody for 1h with shaking. After washing, the reading buffer was added, and plates were read immediately using the MESO QuickPlex SQ 120MM (Meso Scale Discovery, Rockville, MD, USA).

### Data Analysis

Analyses were performed using GraphPad Prism 10.4.1 (GraphPad Inc., San Diego, CA, USA). Initial normality tests indicated that the data were not normally distributed. Consequently, all analyses employed nonparametric tests. Unpaired data were analyzed with the Kruskal-Wallis test followed by Dunn’s multiple comparisons test. Paired data were analyzed using the Wilcoxon signed-rank test. Correlations were assessed using the two-tailed Spearman rank correlation test with a 95% confidence level. All tests were two-sided, and p-values < 0.05 were considered statistically significant. Data are presented as mean ± standard error of the mean (SEM) or geometric mean as specified.

## RESULTS

### Participant Characteristics and biobanking

For this study, 360 participants were recruited at two sites (Quebec City (QC), Canada, and Bobo-Dioulasso (BD), Burkina Faso) and completed two visits spaced 24 ± 2 weeks apart. The visits took place between May 10, 2022, and September 27, 2023, during the predominance of Omicron variants. **Figure 1** presents the schematic design of the study, including description of the study groups (**Fig. 1A**), vaccination coverage across all groups (**Fig. 1B**), and visit timing relative to circulating variants at both locations (**Fig. 1C**).

**Figure 1.**
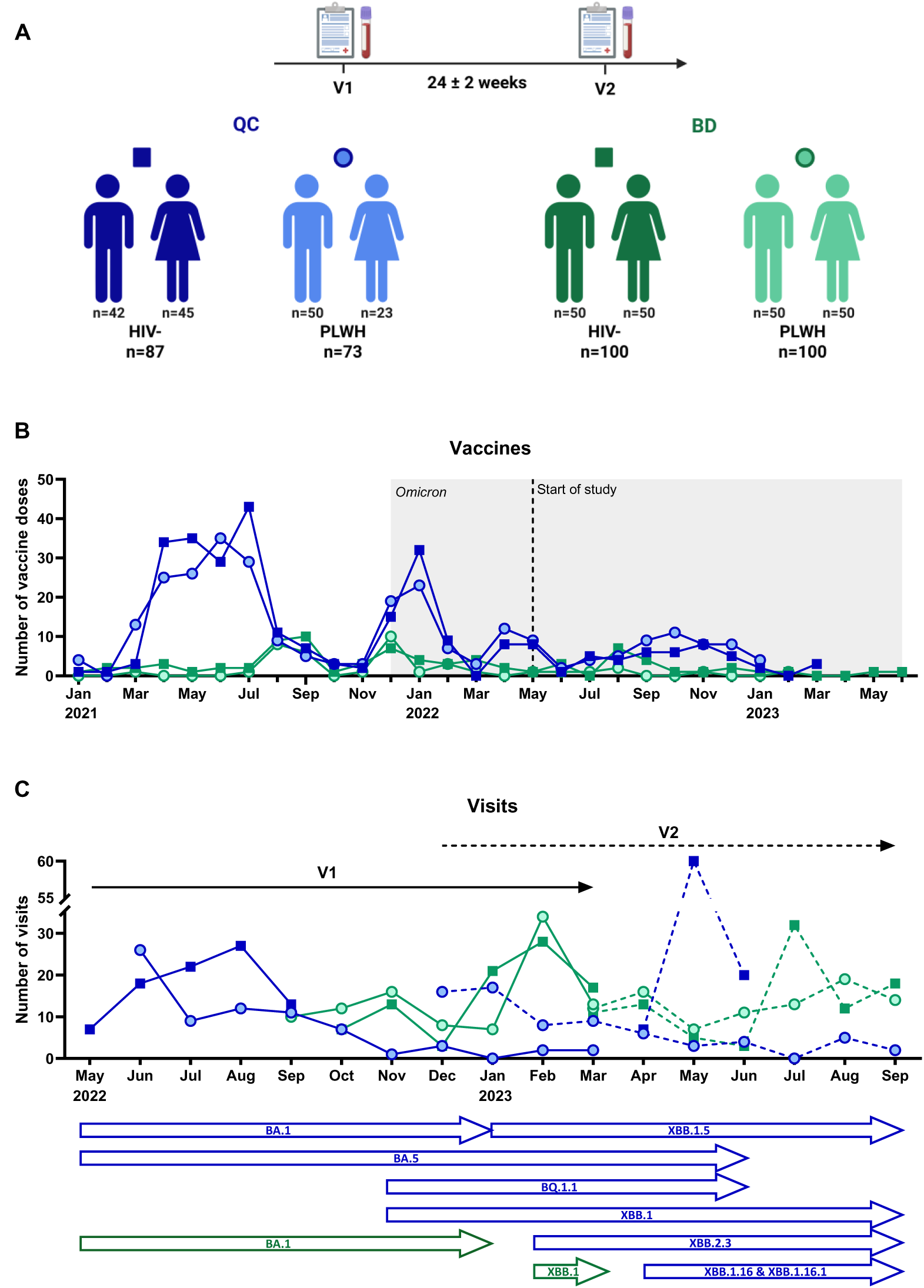
Description of the Study. **(A)** Timeline of the study and group description of cohort participants. Created in BioRender agreement number: YD29OMI2XN). **(B)** Representation of the number of vaccine doses received every month for each group since of the vaccination campaigns. The shaded area indicates the predominance of the Omicron variant beginning at the end of November 2021. **(C)** Representation of the number of participant visits per month for each study group in relation to the circulating variants in QC (blue) and BD (green). QC: Quebec City; BD: Bobo-Dioulasso; HIV-: HIV-negative; PLWH: people living with HIV.

Participants were divided into four groups based on their city of recruitment and their HIV status: 73 PLWH and 87 HIV-participants from QC, and 100 PLWH and 100 HIV-participants from BD (**Table 1**). The study had a 95% retention rate, with 15 participants withdrawing (3 in QC and 12 in BD), and 2 participants in BD who died before the second visit. A total of 168 participants (46.7%) were female and 192 (53.3%) were male; no other sex or gender categories were reported. Participants were distributed as follows: 68 females in QC and 100 in BD, 92 males in QC and 100 in BD, with a median age of 58 (Interquartile range (IQR) 41-64). Among PLWH, 41.1% declared having had COVID-19 since the start of the pandemic in QC and 1.0% in BD, while those numbers were 58.9% in QC and 3.0% in BD for HIV-participants. In the PLWH group, 100% of individuals in QC, and 38.0% in BD received a COVID-19 vaccine, while in the HIV-group, 97.7% and 59.0% were vaccinated, respectively.

**Table 1.**
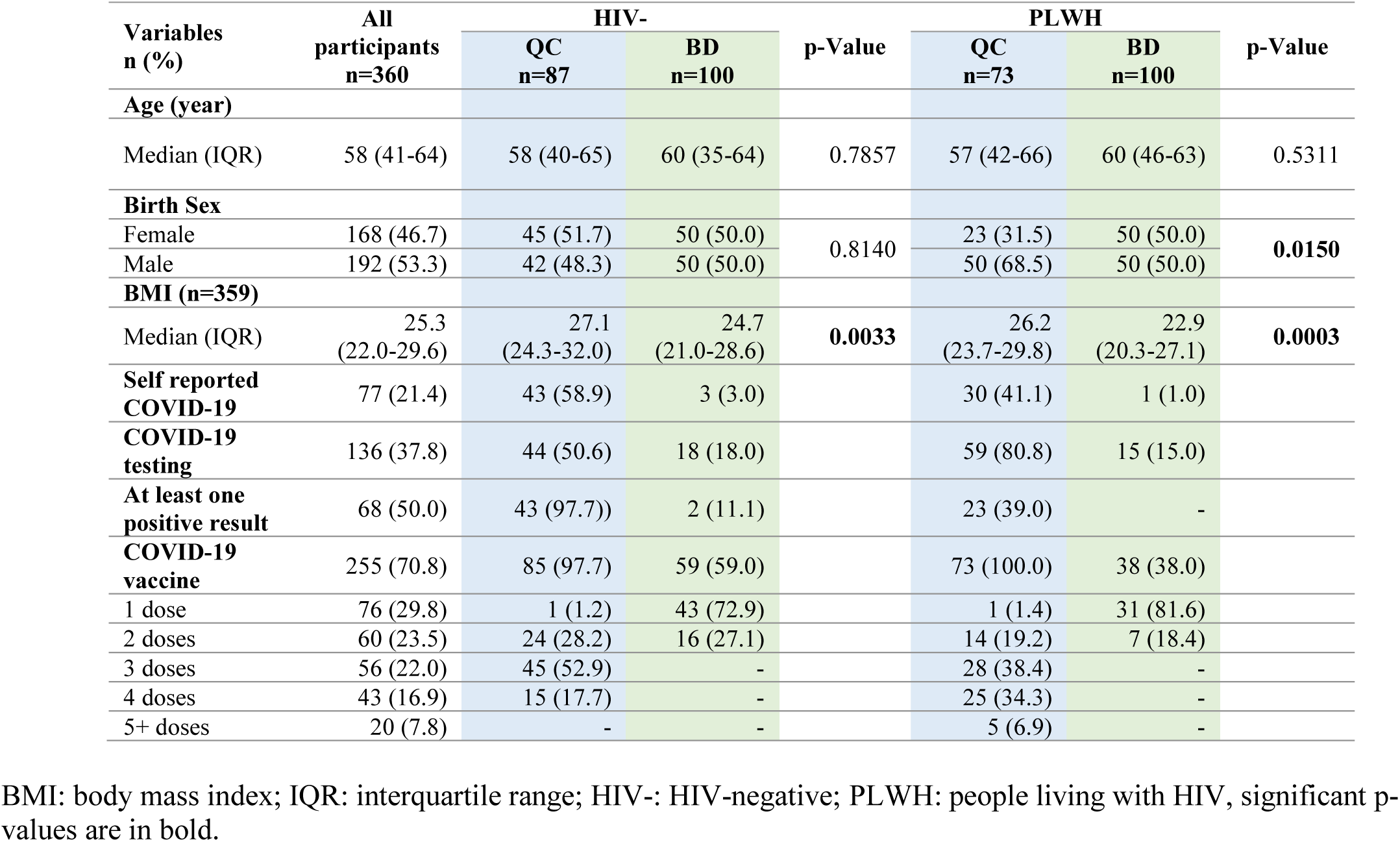
Characteristics of Participants.

The median HIV infection duration in QC was 18.8 years (IQR 12.4-28.9), and 13.3 years (IQR 2.8-18.8) in BD (p< 0.0001), while the median of ART duration was respectively 14.7 years (IQR 9.2-24.8) and 12.1 years (2.8-16.6) (p<0.0001) (**Table 2**). The median CD4 count in QC, 620 cells/μL (IQR 472-854), was comparable to BD, 624 cells/μL (IQR 428-831; p=0.7922). The median CD4 nadir, namely the lowest CD4 count recorded, was 257 cells/μL (IQR 150-369) in QC and 217 cells/μL (IQR 136-347) in BD (p=0.5890). HIV viral load was undetectable for 91.8% of patients in QC and 66.0% in BD (p=0.0002).

**Table 2.** HIV Related Data of Participants.

| Variables n (%) | All participants<br>n=173 | PLWH |  | p-Value |
| --- | --- | --- | --- | --- |
|  |  | QC n=73 | BD n=100 |  |
| **HIV duration (years)** |  |  |  |  |
| Median (IQR) | 14.8 (6.8-20.8) | 18.8 (12.4-28.9) | 13.3 (2.8-18.8) | < **0.0001** |
| **ART** | 172 (99.4) | 73 (100.0) | 99 (99.0) |  |
| **ART duration (years)** |  |  |  |  |
| Median (IQR) | 12.8 (6.3-18.9) | 14.7 (9.2-24.8) | 12.1 (2.8-16.6) | < **0.0001** |
| **CD4 nadir (cells/μL)** |  |  |  |  |
| Median (IQR) | 238 (147-367) | 257 (150-369) | 217 (136-347) | 0.5890 |
| **HIV viral load (n=169)** |  |  |  |  |
| Undetectable (< 20 copies/ml) | 131 (77.1) | 67 (91.8) | 64 (66.0) | **0.0002** |

IQR: interquartile range; ART: antiretroviral therapy; PLWH: people living with HIV. Significant p-values are in bold.

Complete blood counts were done for each participant, as shown in **Figure 2**. Neutrophil counts showed significantly higher numbers in QC compared to BD for both HIV- and PLWH (**Fig. 2A**). CD4^+^ T cells were significantly lower in PLWH in both QC and BD (**Fig. 2B**). In contrast, CD8^+^ T cells were significantly higher in PLWH compared to HIV-as well as in PLWH from QC compared to those from BD (**Fig. 2C**), causing a lower CD4^+^/CD8^+^ ratio in PLWH (**Fig. 2D**). These data indicate differences in CD4^+^ and CD8^+^ T cells counts between HIV-individuals and PLWH, as expected (38). We also observed a difference in neutrophil counts between participants from QC and BD participants consistent with the condition known as benign ethnic neutropenia (BEN) (**Fig.1S**) (39, 40).

**Figure 2.**
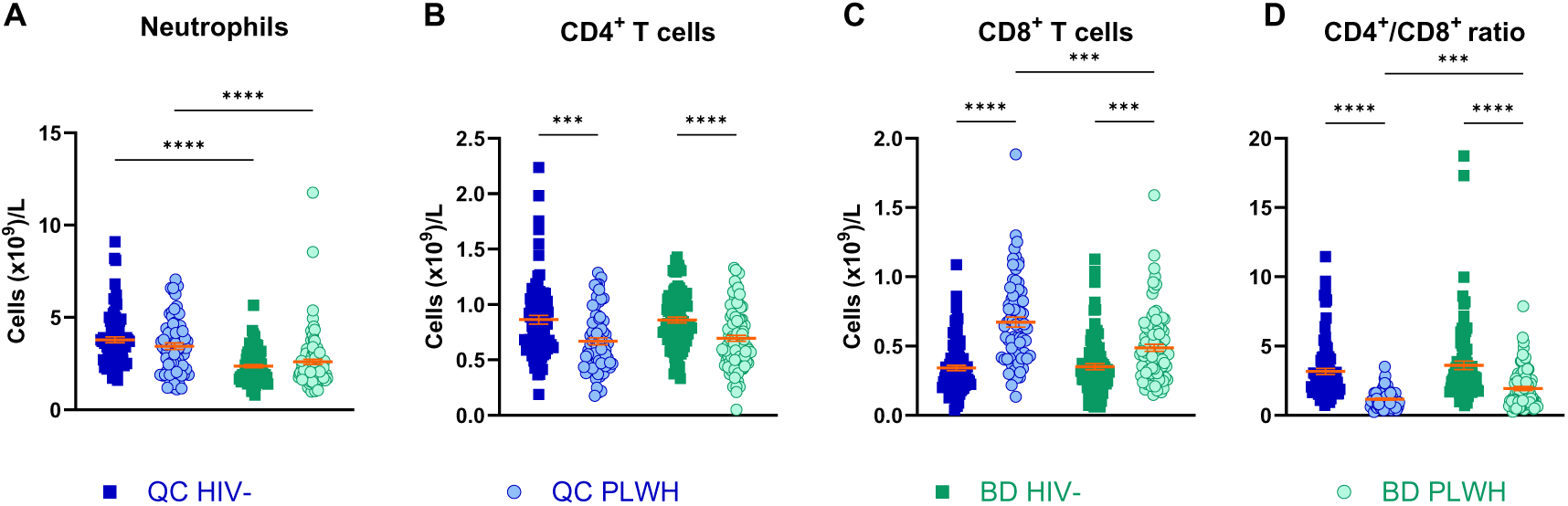
Neutrophil and Lymphocyte Cell Counts. (**A)** Neutrophil count. **(B)** CD4^+^ T lymphocyte count. **(C)** CD8^+^ T lymphocyte ) CD4^+^/CD8^+^ T lymphocyte ratio. QC HIV-n=87, PLWH n=73; BD HIV-n=99, PLWH n=100. Each dot represents a participant. The horizontal solid lines represent the mean ± standard error of the mean. Kruskal-Wallis test, Dunn’s multiple comparisons p < 0.001, **** p < 0.0001.

Regarding the COVID-19 vaccines received, vaccination regimens differed substantially between the two cities. **Table S1** shows the vaccine types and the number of doses received for each group. Available vaccines differed between QC and BD, with mRNA and two-doses viral vector vaccines being available in both locations, but predominantly used in QC, and the one-dose viral vector and inactivated virus vaccines available only in BD. All vaccines have been shown to induce anti-spike (anti-S) antibodies, while inactivated virus vaccines also induce anti-nucleocapsid (anti-N) antibodies (41–46). The majority of participants in QC received monovalent mRNA vaccines targeting the ancestral antigen, while some received bivalent mRNA vaccines targeting either BA.1 or BA.4/BA.5 antigens (17, 47). The primary vaccine given in BD was the one-dose viral vector vaccine.

### Antibody Levels against SARS-CoV-2 Nucleocapsid and Spike Antigens

We measured plasma antibodies against SARS-CoV-2 nucleocapsid (anti-N) and spike (anti-S) antigens across variants to assess the humoral response and whether it differed among them. **Figure S2** shows the determination of the positivity threshold for anti-N and anti-S antibodies as described in the methods section.

### Infection as Determined by Anti-Nucleocapsid Antibodies

Figure 3 shows levels for anti-N antibodies, a marker indicative of prior SARS-CoV-2 infection. Participants who had received inactivated-virus vaccines were excluded from the analysis as these vaccines elicit anti-N antibodies (n=5). At enrollment (Fig. 3A), anti-N antibodies levels were significantly lower in QC when compared to BD. Indeed, less than half (39 %) of the participants in QC and the majority (86 %) in BD had anti-N levels above the positivity threshold (red dotted line), suggesting a higher rate of infection in BD than in QC. At V2 (Fig. 3B), QC participants still had lower titers of anti-N antibodies when compared to BD. However, while the majority of PLWH in QC were still below the threshold, HIV-participants, showed a significant increase in anti-N antibodies. A timeline representation of anti-N titers (**Fig. S3A**) also indicates the increase of cases in HIV-participants from QC, while other groups remain more stable in time.

**Figure 3.**
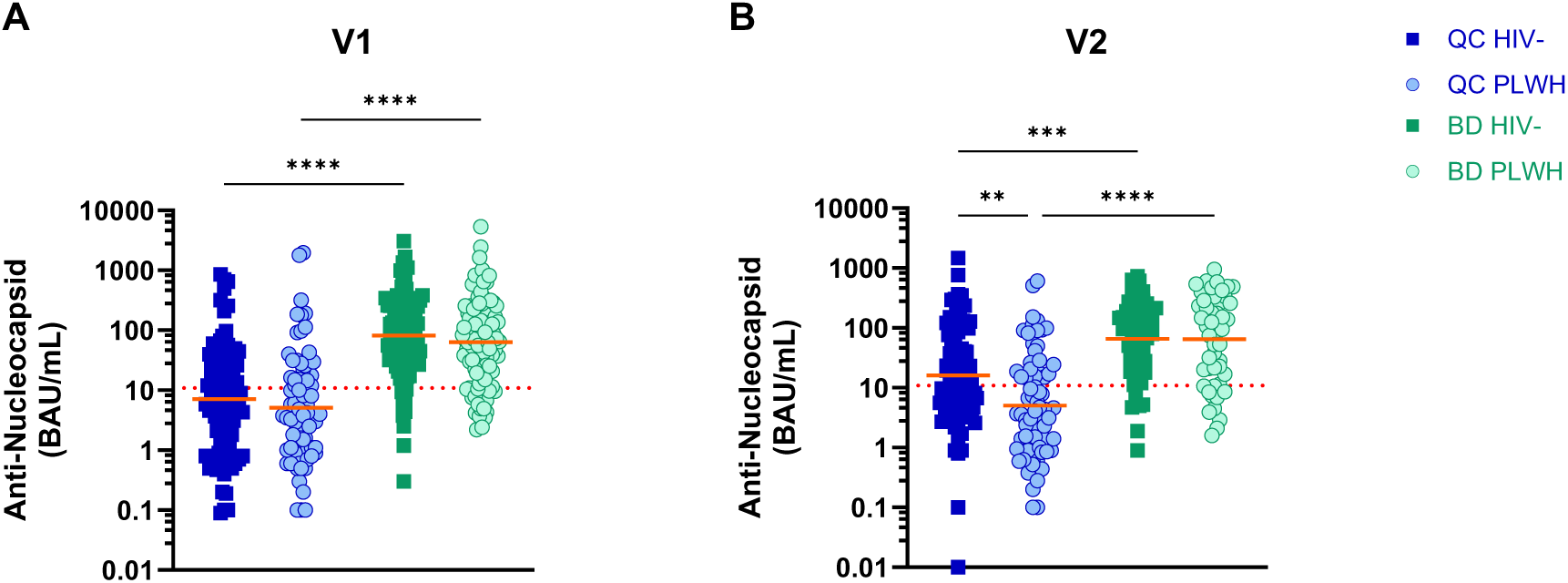
SARS-CoV-2 Anti-Nucleocapsid Antibodies. **(A)** Levels at V1 (QC HIV-n=87, PLWH n=73; BD HIV-n=96, PLWH and **(B)** at V2 (QC HIV-n=87, PLWH n=70; BD HIV-n=56, PLWH n=49). Each dot represents a participant. The dotted line s the positivity threshold. The horizontal solid lines represent the geometric mean. Kruskal-Wallis test, Dunn’s multiple comparisons test. ** p < 0.01, *** p < 0.001, **** p < 0.0001. BAU: Binding Antibody Units.

### Vaccinal and Hybrid Immunity Determined by Anti-Spike Antibodies

Investigation of anti-S (ancestral) antibodies was also conducted to determine vaccinal and hybrid immunity. At enrollment (Fig. 4A), levels of anti-S antibodies were significantly higher in QC than in BD among HIV- and PLWH participants, even though nearly everyone exceeded the positivity threshold. Similar results were observed at V2 (Fig. 4B). These data indicate that most participants were exposed to SARS-CoV-2 infection or vaccination. Again, the serology timeline (**Fig. S3B**) shows that anti-S titers remained relatively stable throughout the study, and that participants from QC had higher titers than those from BD.

**Figure 4.**
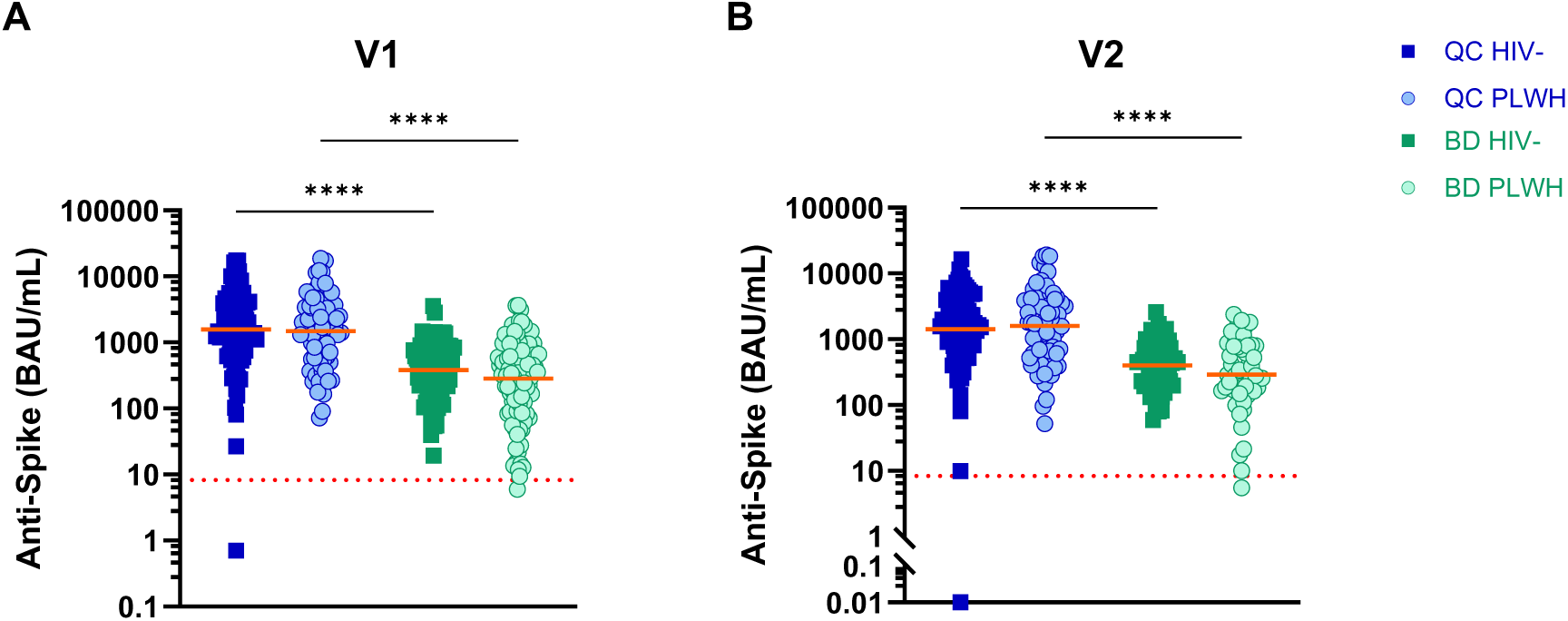
SARS-CoV-2 Anti-Spike (Ancestral) Antibodies. **(A)** Levels at V1 (QC HIV-n=87, PLWH n=73; BD HIV-n=100, PLWH and **(B)** at V2 (24+/- 2 weeks) (QC HIV-n=87, PLWH n=70; BD HIV-n=58, PLWH n=50). Each dot represents a participant. The horizontal solid lines represent the geometric mean. The dotted line represents the positivity threshold. Kruskal-Wallis test, Dunn’s comparisons test. **** p < 0.0001. BAU: Binding Antibody Units.

Correlations were done between the anti-N and anti-S concentrations (**Fig. S4A**). For QC participants, there was no correlation between these two variables (r < 0.3). However, among BD participants, there was a positive correlation (r = 0.5461, p < 0.0001), indicating that anti-S titers increased concurrently with anti-N titers. This suggests that antibody titers may result from infection rather than vaccination. Subsequently, we correlated anti-S titers with the number of days between the visit and the most recent vaccine dose (**Fig. S4B**). Here, we observed a weak negative correlation between the two variables in QC subjects (r = −0.3492, p < 0.0001), indicating that anti-S titers increased after vaccination and then decreased over time. For BD participants, there was no correlation, suggesting that vaccination did not affect antibody levels at the time of the visit, reinforcing the idea that these levels may result from infection.

### Vaccine Type Effect

Since the vaccines administered differed substantially across study groups, we stratified vaccinated participants by vaccine type, either mRNA, viral vector, or inactivated virus vaccines (Fig. 5A). The first type acts by delivering mRNA in lipid nanoparticles to host cells, which produce the antigen and present it to immune cells (48). The viral vector vaccines given to the participants were modified adenovirus used to deliver the target gene (coding for SARS-CoV-2 spike protein) to cells, enabling antigen production and presentation to immune cells (49). The last type is a chemically inactivated whole SARS-CoV-2 virus that still contains all viral components (50). Participants who received a viral vector vaccine have significantly higher anti-N titers than those who received an mRNA vaccine (Fig. 5B). Only five participants received an inactivated virus vaccine, which contains the nucleocapsid antigen; although anti-N titers exceeded the positivity threshold for all participants, it did not reach statistically significant difference. Regarding anti-S titers (Fig. 5C), participants who received an mRNA vaccine had significantly higher titers than the two other groups.

**Figure 5.**
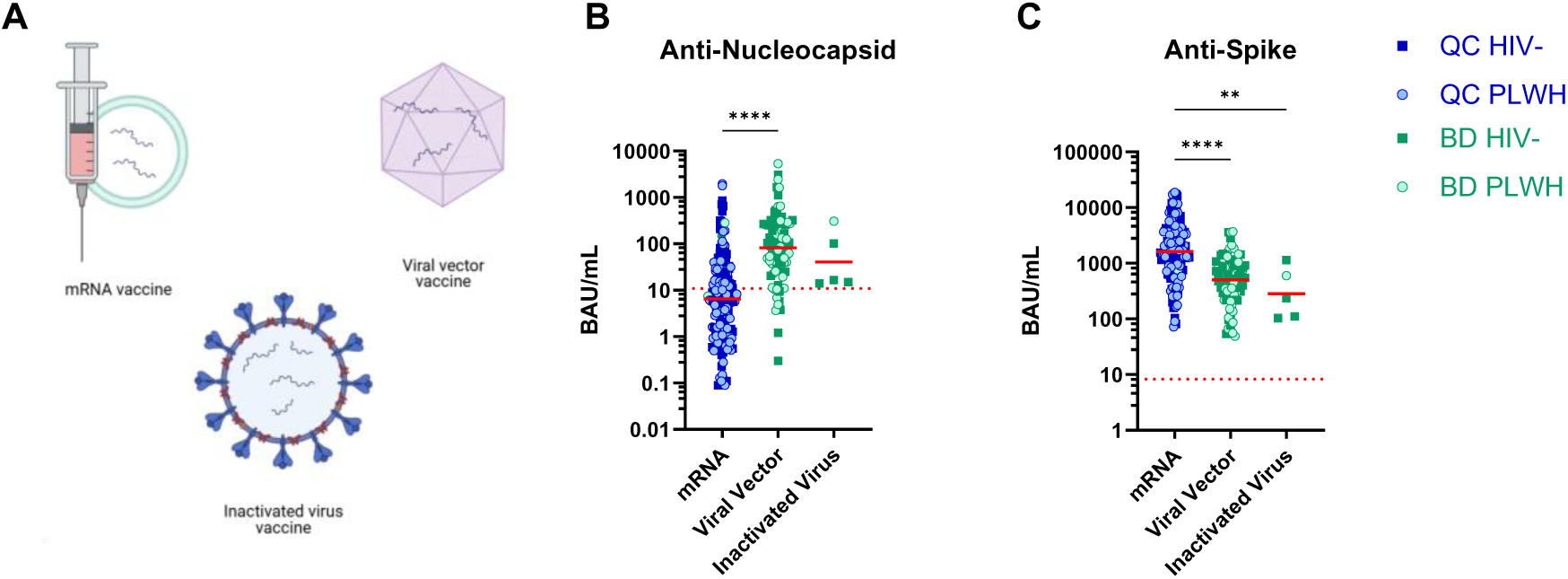
Antibodies Titers by Vaccine Type among Fully Vaccinated Participants. **(A)** Schematic representation of the different vaccines. **(B)** Anti-nucleocapsid antibodies. **(C)** Anti-spike (ancestral) antibodies. mRNA n=161, Viral vector n=88, Attenuated virus n=5. Each dot represents a participant. The horizontal solid lines represent the geometric mean. The dotted line represents the threshold. Kruskal-Wallis test, Dunn’s multiple comparisons test. ** p < 0.01, **** p < 0.0001. BAU: Binding Antibody Units.

### Variants and Bivalent Vaccines

As the study took place during the emergence of the Omicron variant and its multiple subvariants, we compared antibody levels against ancestral spike and nucleocapsid antigens among participants who were unvaccinated, those who received a monovalent vaccine against the ancestral strain, and those who got a bivalent vaccine against the ancestral and Omicron BA.1 or BA.4/BA.5 variants. As shown in Figure 6A, there were significant differences in the titers of ancestral anti-N antibodies among groups, with unvaccinated participants having the highest titers, those vaccinated with monovalent vaccines having intermediate levels, and those vaccinated with bivalent vaccines having the lowest levels. In contrast, for anti-S antibodies (Fig. 6B), non-vaccinated participants had the lowest antibody levels against ancestral spike, whereas those who received monovalent vaccines had intermediate levels, and those who received a bivalent dose had the highest titers. These data suggest that vaccinated participants had a lower infection rate and developed stronger immunity against the spike protein compared to unvaccinated participants. Bivalent vaccines conferred a similar advantage when compared to monovalent vaccines.

**Figure 6.**
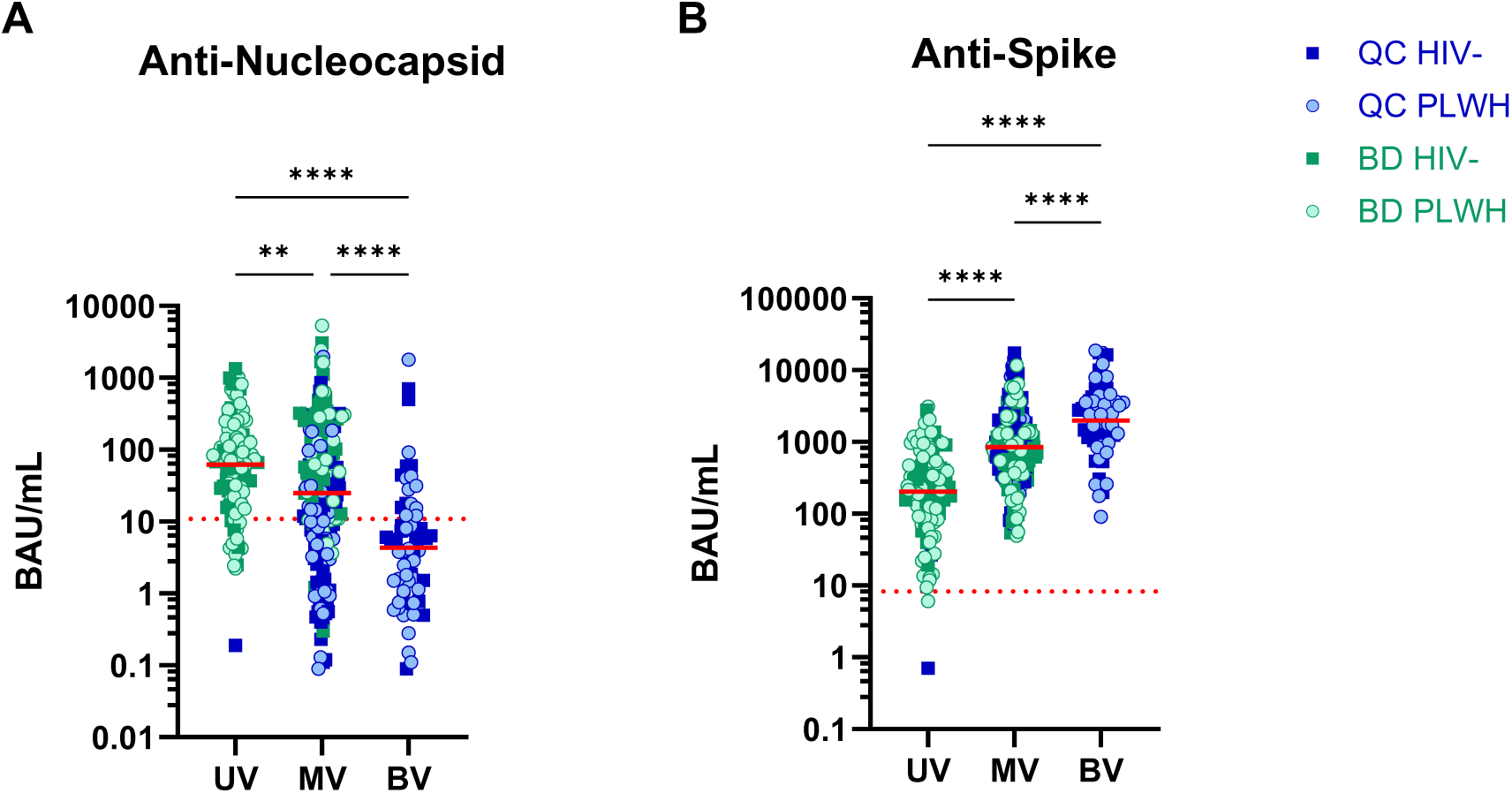
Antibody Titers According to Vaccine Regimen. **(A)** Anti-nucleocapsid antibodies. **(B)** Anti-spike (ancestral) antibodies. represents a participant. The horizontal solid lines represent the geometric mean. The dotted line represents the positivity. Unvaccinated (UV) n=103, monovalent vaccine (MV) n=195, bivalent vaccine (BV) n=62. Kruskal-Wallis test, Dunn’s comparisons test. ** p < 0.01, **** p < 0.0001. BAU: Binding Antibody Units.

Given that antibody levels against ancestral antigens differed significantly between non-vaccinated participants and recipients of monovalent and bivalent vaccines, we sought to determine whether these vaccines induced better antibody response against specific circulating variants. As a reminder, emerging circulating variants during the study period included BA.1, BA.5, BQ.1.1, XBB.1, XBB.1.16, XBB.1.16.1, XBB.1.5, and XBB.2.3.

Figure 7 shows the antibody titers against the spike protein of these different variants. Groups were separated as follows: unvaccinated participants (UV), vaccinated in BD with monovalent vaccines (BD), those who received only monovalent vaccines in QC (MV), and those who received a bivalent vaccine in QC (BV) (no participant in BD received a bivalent vaccine). The results are very similar across all variants. Anti-spike antibody levels were significantly lower in unvaccinated participants than in all other groups for all variants except BA.1, which does not differ between the UV and BD groups. The BD group also has significantly lower antibody levels than both QC groups. However, there are no significant differences between those who received monovalent vaccines and bivalent vaccines in QC.

**Figure 7.**
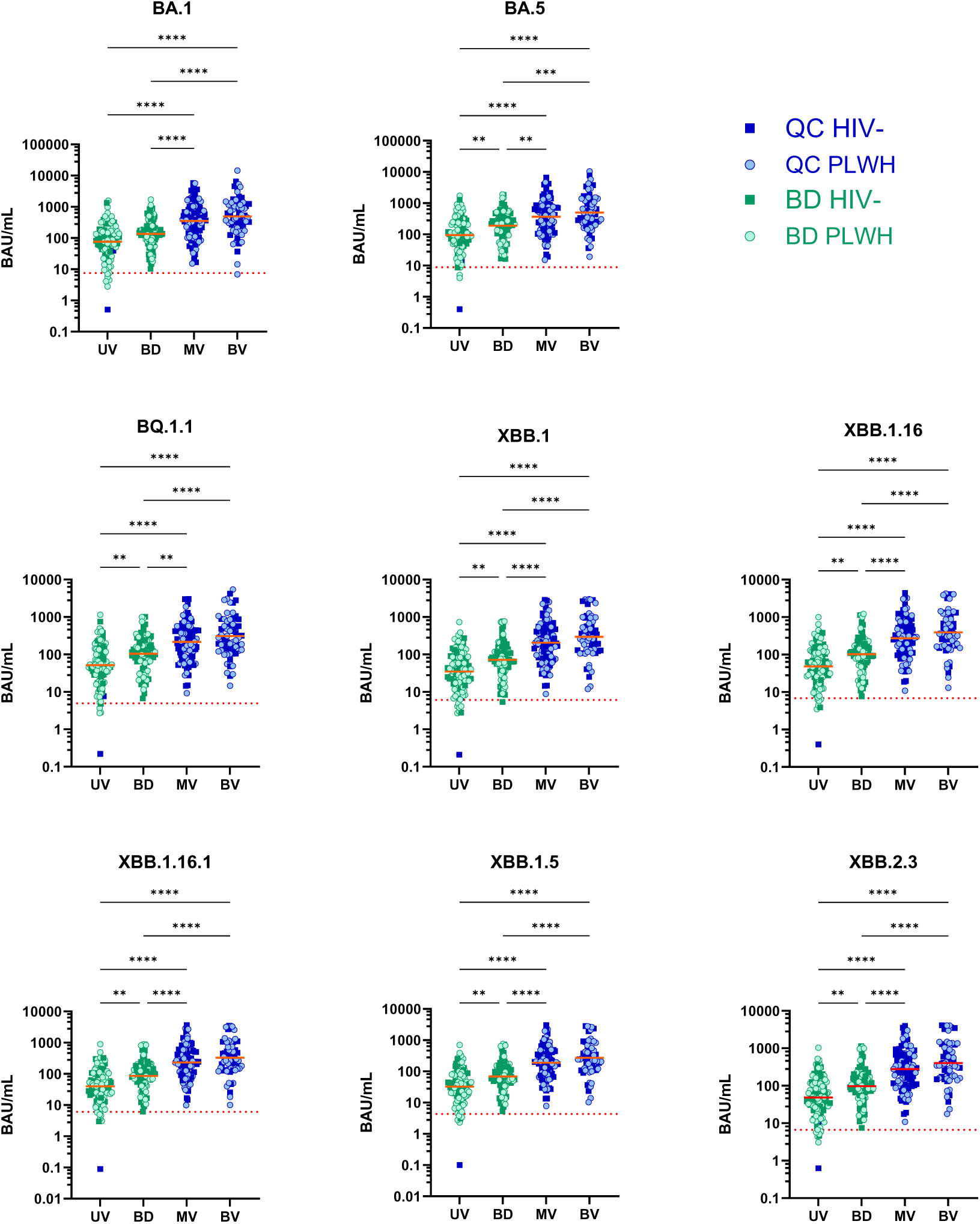
Antibody titers against Spike SARS-CoV-2 Omicron Variants According to Vaccine Regimen. UV: unvaccinated participants (n=105), BD: vaccinated participants in BD (n=97), MV: participants from QC who received a monovalent vaccine (n=96), participants from QC who received a bivalent vaccine (n=62). Each dot represents a participant. The horizontal solid lines represent geometric mean. The dotted line represents the positivity threshold. Kruskal-Wallis test, Dunn’s multiple comparisons test. ** p < p < 0.001, **** p < 0.0001. BAU: Binding Antibody Units.

### Vaccination *versus* Infection

To compare antibody response following vaccination or infection, we reported the percentage of participants with anti-S and anti-N titers above the threshold in those who were vaccinated or reported a SARS-CoV-2 infection in **Table 3**. At enrollment, 98% of HIV- and 100% of PLWH participants in QC were vaccinated, as well as 59% of HIV- and 38% of PLWH volunteers from BD. In QC, 49% of HIV- and 41% of PLWH declared a previous COVID-19, while in BD, only 3% and 1% of HIV- and PLWH, respectively, reported a SARS-CoV-2 infection. In both QC and BD, at least 98% of HIV- and PLWH participants had spike antibody levels above the positivity threshold, confirming the effectiveness of vaccination. In BD, more people had anti-S antibodies than in the vaccinated group, suggesting that a large proportion of those groups were infected. Regarding anti-N antibodies, 43% and 34% of HIV- and PLWH participants in QC, respectively, exceeded the positivity threshold, consistent with the self-assessment results. In BD, however, 91% of HIV- and 82% of PLWH participants were above the positivity threshold for anti-N antibodies, supporting a much greater rate of infection than reported by the self-assessment. For all parameters: vaccination status, positivity for anti-S and anti-N antibodies, and COVID-19 self-assessment, V2 data were similar to those of V1 across all groups, except for the COVID-19 self-assessment among QC participants, where a higher proportion of HIV-participants and lower proportion of PLWH reported an infection than at V1.

**Table 3.** Comparison between Markers of SARS-CoV-2 Infection or Vaccination and Self-Assessment.

|  |  | Vaccination |  | Self-reported COVID-19 infection |  | Spike antibodies |  | Nucleocapsid antibodies |  | BD infection test |  |
| --- | --- | --- | --- | --- | --- | --- | --- | --- | --- | --- | --- |
|  |  | V1 | V2 | V1 | V2 | V1 | V2 | V1 | V2 | V1 | V2 |
|  |  | % |  |  |  |  |  |  |  |  |  |
| QC | HIV- | 98 | 98 | 49 | 75 | 99 | 99 | 43 | 54 | - | - |
|  | PLWH | 100 | 100 | 41 | 9 | 100 | 100 | 34 | 34 | - | - |
| BD | HIV- | 59 | 61 | 3 | 0 | 100 | 100 | 91 | 90 | 43 | 24 |
|  | PLWH | 38 | 38 | 1 | 0 | 99 | 98 | 82 | 78 | 34 | 22 |

QC: Quebec City; BD: Bobo-Dioulasso; HIV-: HIV-negative; PLWH: people living with HIV.

## DISCUSSION

This observational study aimed to investigate antibody responses in PLWH following SARS-CoV-2 infection and vaccination across two populations BD and QC, and to compare them to HIV-participants in each region. The volunteers were sampled at two visits spaced 24 weeks apart. At baseline, the four groups were similar in age. PLWH in both centers had comparable CD4 counts and nadirs. However, PLWH from QC had a longer duration of known seropositivity and ART treatment, and their viral load was more often undetectable.

No difference in anti-S antibody titers was observed between HIV-participants and PLWH at both enrollment cities and at V2. Anti-N antibodies were also similar between PLWH and HIV-participants except at V2 in QC where HIV-participants had significantly more antibodies than PLWH, consistent with the PLWH group’s self-assessments reporting fewer infections. This difference may be due to better compliance with COVID-19 public health measures among PLWH in QC than among control volunteers. Furthermore, infection rates in BD were substantially higher than those reported by self-assessments. This confirms reports that there were not fewer cases in sub-Saharan Africa; instead, there might have been more asymptomatic cases and fewer detections due to poorer screening (3, 4). Additionally, the significantly higher levels of anti-N antibodies suggest that participants in BD were infected more recently than those in QC, as antibody levels tend to decline with time (51, 52). Moreover, the detection of anti-S antibodies in nearly all participants indicates that the entire study group was exposed to the antigen, either through vaccination or viral infection. Participants from QC were more vaccinated, with many receiving more than 3 doses; consequently, they had higher anti-S antibody levels than volunteers from BD. Indeed, antibody titers increase with each vaccine dose received, as shown by other groups (53, 54). More than half of the participants in BD remained unvaccinated throughout the study, indicating that they developed antibodies primarily through SARS-CoV-2 infection. It is also supported by the positive correlation between anti-S and anti-N antibodies among participants with BD.

This study also showed that vaccine regimens appeared to affect the antibody titers against both spike and nucleocapsid proteins. Although establishing a cause-and-effect relationship is difficult given the many variables involved, we observed that participants with higher anti-S antibody titers also had lower anti-N titers, suggesting a less severe SARS-CoV-2 infection among well-vaccinated participants. Moreover, the types of vaccines administered (mRNA, viral vector, or inactivated virus) induced significantly different antibody titers against the spike proteins. These differences could represent the number of doses received rather than being related to the type the vaccines. Indeed, most participants who received mRNA vaccines received booster doses in QC, meaning they had more doses and less time elapsed between the last vaccination and the study visit. This is also supported by the higher anti-S titers observed in individuals who received a bivalent vaccine (available at a later time point). By contrast, those who were vaccinated with viral vector or inactivated virus vaccines have mostly received only one or two doses at short intervals, leaving more time before the visit and resulting in a more pronounced decline in antibodies.

### Strengths and Limitations

We conducted an observational study of 360 PLWH and HIV-participants across two regions and populations, enabling us to study humoral and potentially cellular innate and acquired immune responses in each subgroup relative to the SARS-CoV-2 virus. In addition, the clinical information collected at both visits provided essential data to correlate clinical parameters with experimental results. We have efficiently transferred experimental methods between laboratories to obtain comparable samples. Moreover, we achieved a high subject retention rate (95%), yielding two measures for most participants.

Among the study limitations is its timing, which occurred relatively late in the pandemic, when most participants had been exposed to the SARS-CoV-2 antigens through vaccination or infection, thereby precluding the establishment of a naïve group. Furthermore, vaccination data in BD were collected based only on participants’ memories, introducing a recall bias. We also cannot exclude a social desirability bias that can lead some participants to report that they were vaccinated even though they were not. Multiple variables had to be considered, which complicated the result analysis, such as the different vaccines available in QC and BD, differences in the number of doses received between the two locations, and differences in the time elapsed between vaccination and visits. Moreover, the positivity threshold has been established using QC samples exclusively, potentially introducing a bias in the analysis of BD samples. A few studies have examined cross-reactivity in pre-pandemic samples and found that a non-negligible proportion of the population in sub-Saharan Africa had antibodies that bound the SARS-CoV-2 spike and nucleocapsid proteins, a phenomenon not observed in Western populations (55–57). Therefore, this might indicate that the number of infections in the BD population is overestimated in our study.

### Perspectives

With more than 700 samples of diverse blood components, we can study multiple aspects of immunity across populations and examine how they are affected by SARS-CoV-2 vaccination and infection. It is also possible to study neutrophil responses as part of trained immunity in relation to vaccines and SARS-CoV-2 infection. This will enable us to produce evidence that contributes to a better understanding of vaccine and infection-induced responses in individuals with immune deficiencies. We have also collected hundreds of PBMC samples for studying the adaptive cellular response using a proliferation assay, a protocol already established in our laboratory. With the plasma samples collected, we intend to study the different functional antibodies elicited by vaccination and infection.

In addition, this approach allows studying and comparing the innate response between the two locations. Indeed, it is suggested that the population in Africa has better-trained immunity due to multiple factors, such as a higher Bacille Calmette-Guérin vaccination rate and more frequent epidemics, which could explain in part why the pandemic had a lesser impact on that continent (4). Thus, examining the innate response may help predict which individuals are most likely to mount optimal responses to vaccines or viruses in terms of intensity and duration.

### Conclusion

This study shows that treated PLWH mount robust humoral immune response to SARS-CoV-2 vaccination and infection, comparable to HIV-controls. We found that these two populations of treated PLWH were not more susceptible to contracting COVID-19 than HIV-individuals. Instead, the differences observed were attributable to participants’ geographic locations with respect to vaccination and infection: participants in QC were more vaccinated and thus had stronger humoral immunity to the spike protein, whereas those in BD had more SARS-CoV-2 infections, as indicated by higher anti-N antibody titers.

## Acknowledgements

The authors thank the research coordinator, Isabelle Chabot and the nurses Dany Poulin, Geneviève Corneau, Marie-Christine Samson and Geneviève Gagnon at UHRESS *(“Centre de recherche du CHU de Québec-Université Laval”*), the nurse Assane Savadogo, the lab technicians Alidou Zango, Roland Gnoumou, Moumini Nouctara, Clement Sanou, and Sedina Traore at the Centre Muraz laboratory and Yerelon Clinic for their involvement in this study. We are very grateful to the study participants, without whom this study would not have been feasible. The authors thank the *“Plateforme de Bio-fabrication de Vaccins Québec/Centre de Recherche du CHU de Québec-Université Laval”* for the MESO^®^ QuickPlex SQ 120MM service.

## Author Contributions

HJ: Conceptualization, Data curation, Formal analysis, Investigation, Methodology, Writing – original draft, Writing – review & editing. WWB: Funding acquisition, Project administration, Supervision, Data curation, Formal analysis, Investigation, Methodology, Writing – review & editing. IB, AP: Investigation, Methodology, Writing – review & editing. JV: Investigation, review & editing. MT, ST, ITT, DK: Resources, Writing – review & editing. ATO; YZ: Investigation, Supervision, Writing – review & editing. MP, PAT, MP, MLV: Funding acquisition, Resources, Supervision, Validation, Writing – review & editing. CG: Conceptualization, Funding acquisition, Project administration, Investigation, Formal analysis, Resources, Supervision, Writing – review & editing.

## Ethical Statement

The study was approved by the ethics committee of the “*Centre de recherche du CHU de Québec-Université Laval”* (registration number CER-2022-6241, CER-2023-6445, CER-2021-5744, CER-2019-4163, CER12-03-167) and Burkina Faso Ministry of Health Research Ethics Committee (Deliberation n°2022-03-048).

## Data availability statement

The study protocol, results and informed consent documents will be made available to researchers upon request from the corresponding author. Researchers will be asked to complete a concept sheet for their proposed analyses to be reviewed, and the investigators will consider the overlap of the proposed project with active or planned analyses and the appropriateness of the study data for the proposed analysis.

