## Supplementaldata for "SARS-CoV-2 Antibody Response during Omicron Predominance after COVID-19 Vaccination in People Living with HIV: A Comparative Study in Canada and Burkina Faso": COVID-VIH_Supplementary file_2026-05-25(JVirology).pdf

### Supplementary Material

**Table S1. Number of Vaccine Doses Received by All Participants for Each Available Vaccine.**

| | | Pfizer | | Moderna | | AstraZeneca | Johnson & Johnson | Sinopharm | Sinovac |
|---|---|---|---|---|---|---|---|---|---|
| | | M* | B** | M* | B** | | | | |
| QC | HIV- | 108 | 11 | 116 | 16 | 26 | - | - | - |
| | PLWH | 133 | 24 | 86 | 13 | 17 | - | - | - |
| BD | HIV- | 4 | - | 1 | - | 21 | 42 | 6 | 1 |
| | PLWH | 4 | - | 1 | - | 9 | 29 | 2 | - |

*Monovalent, **Bivalent, HIV-: HIV-negative; PLWH: people living with HIV.

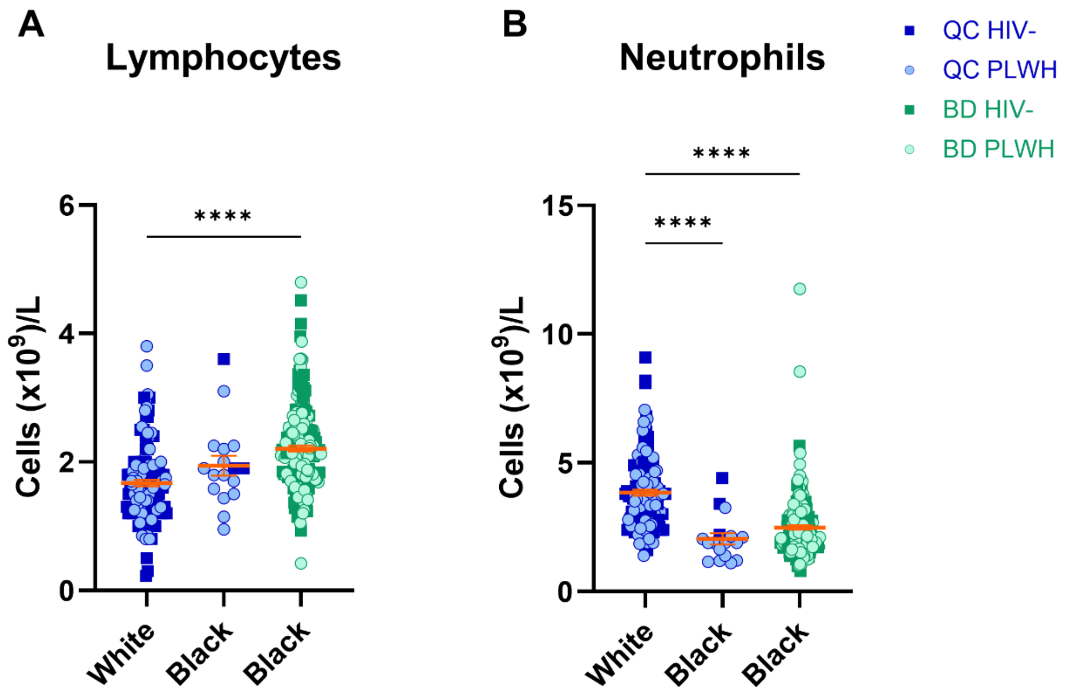


**Supplementary Figure 1. Data comparison in pooled PLWH and HIV- participants between ethnicities. (A)** Lymphocyte count. **(B)** Neutrophil count. White QC n=136, Black QC n=17, Black BD n=198. Kruskal-Wallis test, Dunn's multiple comparisons test. Data are shown as mean ± SEM. **** $p < 0.0001$

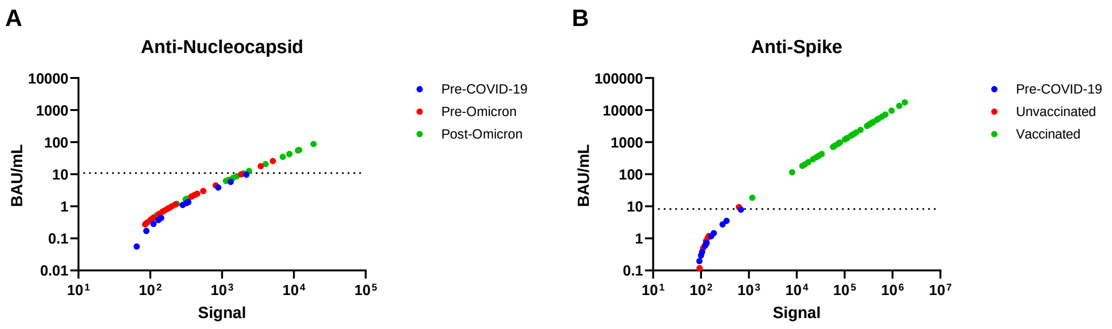


**Supplementary Figure 2. Positivity Threshold Determination. (A)** Anti-nucleocapsid antibodies. **(B)** Anti-spike antibodies. The signal was measured for pre-COVID-19 plasma samples (blue), and the mean was calculated; the threshold was set at three standard deviations above the mean. Pre- and post-omicron plasma samples, some from vaccinated individuals and others from unvaccinated individuals, were used as controls. BAU: Binding Antibody Units.

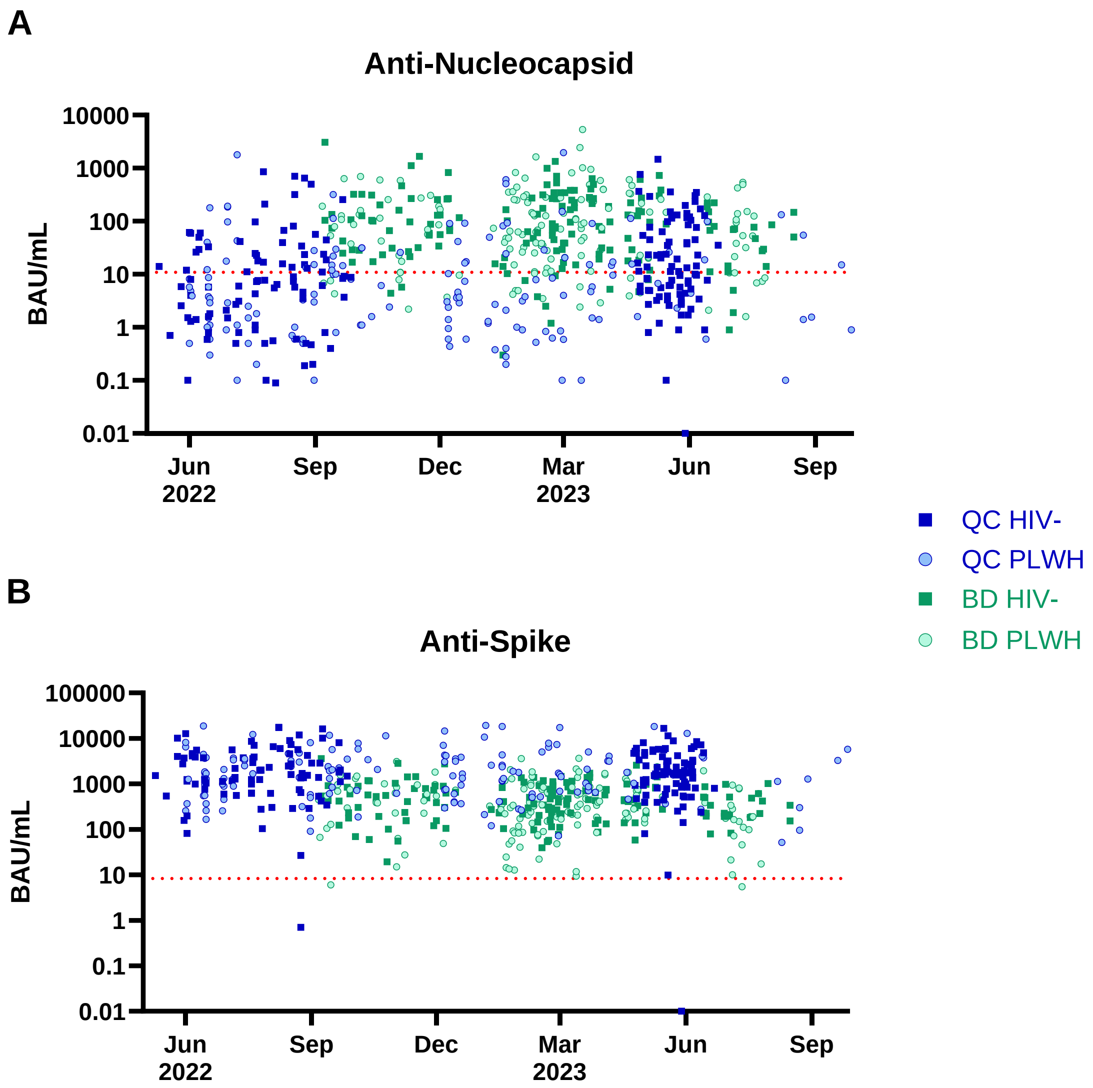


**Supplementary Figure 3. Serology Timeline. (A)** Anti-Nucleocapsid and **(B)** anti-Spike titers of each participant during visits throughout the study. BAU: Binding Antibody Units.

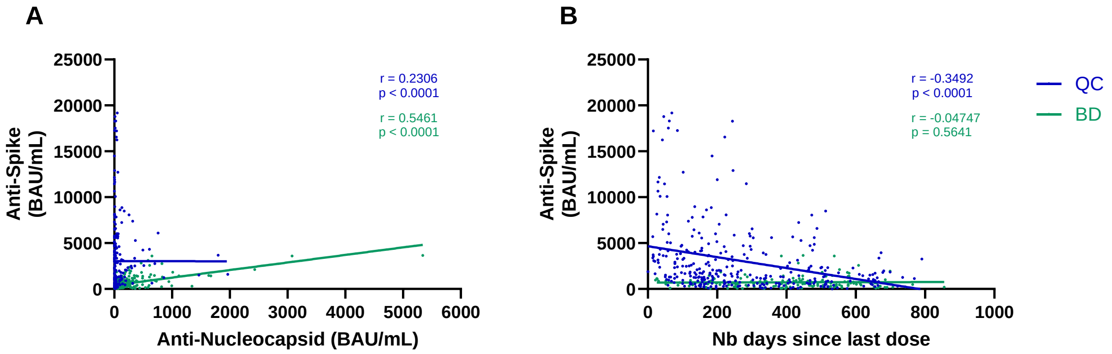


**Supplementary Figure 4. Anti-Spike Titer Correlations. (A)** Correlation with anti-Nucleocapsid titers. QC n=317, BD n=308. **(B)** Correlation with the number of days between the visit and the last vaccine dose received. QC n=313, BD n=150. Blue dots represent participants from QC, while green dots represent BD participants' data from V1 and V2. A two-tailed Spearman correlation with a 95% confidence interval was used. BAU: Binding Antibody Units.

#### Appendix I – Blood Samples Processing Protocol

Sampling:

- 2 x 2.7 mL-citrate tubes
- 2 x 6 mL-serum tubes
- 2 x 4 mL-heparin tubes
- 9 x 10 mL-EDTA tubes

Immediately after sampling, anticoagulant-containing tubes were placed on a rotating platform at room temperature (RT). Processing was started within two hours of sampling.

Solutions:

*HEPES 1M*

1. Weigh 119.16 g HEPES (Wisent, Cat# 600-032-CG).
2. Add 350 mL distilled water.
3. When completely dissolved, adjust pH to 7.4 with NaOH.
4. Complete volume at 500 mL.
5. Filter at 0.22 µm into a sterile bottle.
6. Store at 4°C.

*$CaCl_2$ 0.16M*

1. Weigh 4.7 g $CaCl_2$•$2H_2O$ (Fisher, Cat# C79-500).
2. Add 150 mL distilled water.
3. Dissolve with a magnetic stirrer.
4. Complete volume to 200 mL and mix well.
5. Filter at 0.22 µm into a sterile bottle.
6. Store at 4°C.

*2% Dextran*

1. Weigh 10 g dextran (Sigma, Cat# 31392-50G) and transfer to a beaker.
2. Add 50 mL HBSS 10X (Wisent, Cat# 311-506-CL), 5 mL $CaCl_2$ 0.16M and 5 mL HEPES 1M.
3. Add distilled water up to around 400 mL.
4. Dissolve with a magnetic stirrer until homogeneous.
5. Adjust pH to 7.4 with NaOH.
6. Complete volume to 500 mL.
7. Filter at 0.22 µm into a sterile bottle.
8. Store at 4°C.

*HBSS 1X for Neutrophil Isolation*

1. Add 100 mL HBSS 10X (Wisent, Cat# 311-506-CL; PAN Biotech, Cat# P04-34500), 10 mL HEPES 1M and 10 mL $CaCl_2$ 0.16M into a beaker.
2. Add distilled water to a volume of 900 mL.

3. Adjust pH to 7.4 with NaOH.
4. Complete final volume to 1 L and mix well.
5. Filter at 0.22 µm into a sterile bottle.
6. Store at 4°C.

*Trypan Blue*

1. Dilute 5 mL trypan blue (Wisent, Cat# 609-130-EL) with 5 mL HBSS 1X (Wisent, Cat# 311-512-CS).
2. Filter at 0.22 µm into a sterile 15 mL tube.
3. Store at RT.

*Decomplemented Fetal Bovine Serum (FBS)*

1. If the serum bottle (Sigma, Cat# F1051-500ML; PAN Biotech Cat# P30-5500) is kept at -20°C, thaw at 4°C.
2. When the FBS is completely thawed, mix by inversion 2-3 times.
3. Place in a shaking waterbath with the water level covering the liquid in the bottle.
4. Stabilize the bottle with a heavy-weight ring.
5. When the temperature reaches 56°C, mix by inversion 2-3 times every 10 min.
6. Let the bottle sit in the water bath for 30 min.
7. Quickly cool the bottle by immersing it in a container of iced water up to the liquid level.
8. Aliquot into 50 mL tubes under a hood.
9. Seal with Parafilm and store immediately at -20°C.

*Freezing Medium 2X*

1. Mix 2 mL DMSO (Sigma, Cat# D2650-5X10ML) with 8 mL decomplemented FBS (Sigma, Cat# F1051-500ML; PAN Biotech Cat# P30-5500).
2. Store at 4°C.

*Complete RPMI (2% FBS)*

1. Add 10 mL decomplemented serum to a 500 mL bottle of RPMI (Wisent, Cat# 350-000-CS).
2. Mix the bottle by inverting it a few times.

**Protocol:**

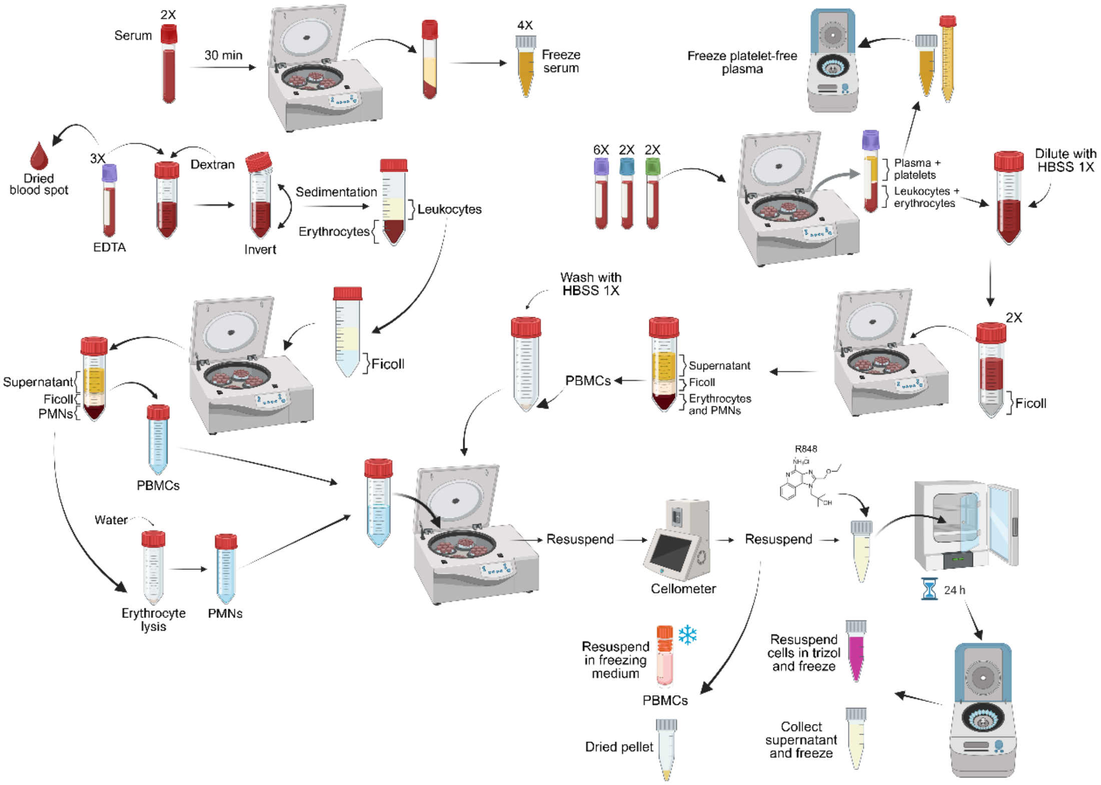


**Supplementary Figure S5. Blood Separation Protocol.** After sampling, the serum tubes are allowed to rest to facilitate clotting, then centrifuged to collect serum. A few blood droplets are taken from an EDTA tube to make a dried blood spot. Three EDTA tubes are incubated with 2% dextran for 30 min to allow erythrocytes to sediment. The supernatant left is used to isolate PBMCs and PMNs. Meanwhile, the citrate, heparin, and EDTA tubes left are centrifuged to separate plasma. EDTA plasma is collected in a 15 mL tube, centrifuged, and frozen. Citrate, heparin, and EDTA plasma are aliquoted in 1 mL and centrifuged to remove platelets before freezing. The remaining blood cells are diluted in HBSS and used to isolate PBMCs. The PBMC ring is collected from all tubes and washed. Isolated cells are counted and separated for R848 stimulation or freezing. Created in BioRender (agreement number: EF299C0RUK).

*All solutions must be used at room temperature, unless stated otherwise.

*Serum Preparation*

1. Let serum tubes rest vertically, without rotating, for at least 30 min after sampling.
2. Centrifuge tubes at 1600 *g* for 15 min without breaks.
3. Collect 2 x 1 mL serum samples per 1.5 mL tube.
4. Freeze at -20°C and transfer at -80°C for long-term storage.

*DBS Preparation*

1. Pipet 5 x 80 µL from an EDTA tube and gently place on the circles of the filter paper (Sigma, Cat# WHA10534612; Cytiva, Cat# 903 Protein saver card).
2. Let dry under the hood for approximately 3 hours.

3. Place in a sealable plastic bag with desiccant bags (Fisher, Cat# 09-928-142) and glycine (Fisher, Cat# 09-924-203) paper separating the DBS cards. Add humidity indicators (Fisher, Cat# AC448430250), seal and store at -20°C.

*Plasma*

1. Combine the citrate and heparin tubes into two separate 15 mL tubes.
2. Centrifuge citrate, heparin, and six EDTA tubes at 500 *g*, acceleration 9 (A9) and deceleration 5 (D5), for 10 min.
3. Collect 15 mL of EDTA plasma into a 15 mL tube and centrifuge at 600 *g* for 10 min. Dispose of the remaining plasma in the liquid waste.
4. Collect 2 x 1 mL centrifuged EDTA plasma samples into 1.5 mL tubes and transfer the remaining 13 mL into a new 15 mL tube, avoiding the pellet.
5. Collect 2 x 1 mL aliquots of citrate and heparin-plasma into 1.5 mL tubes.
6. Centrifuge the six tubes at 3000 *g* for 10 min.
7. Transfer the plasma into new 1.5 mL tubes, ensuring that the platelet pellet is not transferred.
8. Freeze all plasma at -20°C, then transfer to -80°C for long-term storage.

*PBMC and PMN Isolation for R848 Activation*

1. Transfer the whole blood from three EDTA tubes into a 50 mL tube, add 10 mL of 2% dextran solution, and gently mix by inverting.
2. Loosen the cap and allow the sediment to settle for up to 30 min.
3. Collect the supernatant and place it on top of 15 mL lymphocyte separation medium (Ficoll) (Wisent, Cat# 305-010-CL; PAN Biotech, Cat# P04-66500).
4. Centrifuge Ficoll tubes at 600 *g* for 20 min, A5/D2.
5. Collect the PBMC ring and transfer the remaining supernatant to the liquid waste.
6. Tap on the tube to resuspend the PMN pellet.
7. Add 36 mL of sterile milli-Q water to the PMN pellet to remove contaminating red blood cells, then transfer the suspension to a 50 mL tube containing 4 mL HBSS 10X (Wisent, Cat# 311-506-CL) to stop the lysis.

*PBMC (EDTA) Isolation for Biobanking*

1. Combine the six EDTA tubes without plasma in a 50 mL tube and bring the volume to 40 mL with HBSS 1X.
2. Mix and collect 2 x 20 mL samples, then place them on top of 15 mL lymphocyte separation medium in two separate 50 mL tubes.
3. Centrifuge Ficoll tubes at 600 *g* for 20 min, A5/D2.
4. Collect the PBMC ring in 50 mL tubes and discard the remaining cell pellet in the solid waste bag.
5. Wash PBMCs by adding HBSS 1X to the 40 mL line.

*PBMC (Citrate and Heparin) Isolation for Other Uses*

1. Complete the citrate and heparin tubes to 6 mL with HBSS 1X, then transfer the contents onto the surface of 6 mL of lymphocyte separation medium in two 15 mL tubes.
2. Centrifuge Ficoll tubes at 600 *g* for 20 min, A5/D2.

3. Collect the PBMC ring in 15 mL tubes and discard the remaining cell pellet into the solid waste bag.
4. Wash PBMCs by adding HBSS 1X to the 10 mL line.

*Cell Count*

1. Centrifuge all tubes at 400 *g* for 10 min.
2. Remove the supernatant and resuspend the EDTA- and dextran-PBMCs, as well as PMNs, in 5 mL of HBSS 1X for cell count; transfer the cells to new 15 mL tubes. Resuspend citrate and heparin PBMCs in 1 mL HBSS 1X.
3. Count cells (EDTA and dextran PBMCs, and PMNs) by diluting 20 µL of cells with 20 µL of diluted trypan blue using the Cellometer Vision cell counter (Nexcelom Bioscience LLC, Lawrence, MA, USA).

*Cell Stimulation with R848*

1. Centrifuge dextran PBMCs and PMNs at 400 *g* for 5 min.
2. Resuspend at 20 x $10^6$ cells/mL with X-Vivo medium (Lonza Bioscience, Cat# BEBP02-061Q).
3. Identify 12 eppendorf tubes (1.5 mL): 6 for PBMCs and PMNs in the resting condition 1h, 4h or 24h, and 6 for R848 activation.
4. Place 1 µL of R848 (Cedarlane Laboratories, Cat# VAC-R848) at 1 mg/mL (final conc. 1 µg/mL) into the 6 activation tubes.
5. Add 750 µL X-Vivo medium in all PMN tubes (Resting and R848) and 875 µL in all PBMC tubes.
6. Add 250 µL of PMNs (5 x $10^6$ cells) into all six tubes, and 125 µL of PBMCs (2.5 x $10^6$ cells) into the six tubes.
7. Loosen the cap and incubate at 37°C + 5% $CO_2$ for the specified time.
8. Centrifuge the tubes at 400 *g* for 5 min.
9. Collect the supernatant and transfer it into a new Eppendorf tube.
10. Resuspend the cell pellets with 1 mL of cold (4°C) TRIzol Reagent (Fisher, Cat# 15596018).
11. Freeze the supernatant and TRIzol tubes at -20°C, then transfer them to -80°C for long-term storage.

*Freezing PBMC [1]*[1]

1. Centrifuge EDTA, citrate, and heparin PBMC at 400 *g* for 5 min.
2. Aspirate supernatant and resuspend citrate and heparin PBMCs with 500 µL of cold (4°C) decomplemented FBS, then transfer to cryovials.
3. Resuspend EDTA PBMCs at 40 x $10^6$ cells/mL according to the cell count with cold decomplemented FBS, and transfer 500 µL per cryovial (20 x $10^6$ cells/tube).
4. Using the vortex at a low setting, add 500 µL (or a 1:1 volume) of cold 2X freezing medium dropwise to each cryovial to obtain a total volume of 1 mL.
5. Place quickly in a cold (4°C) freezing block and transfer to -80°C for 24h.
6. Transfer cells in a cryobox at -150°C for storage.

[1] Adapted from BQC19

*Dry Pellets*

1. If dextran PBMCs and PMNs remain, transfer 5 x $10^6$ PMNs (250 µL) and 2.5 x $10^6$ dextran PBMCs (125 µL) into 1.5 mL tubes (1 PMN and 2 PBMC tubes).
2. Centrifuge 5 min at 400 *g*.
3. Remove the supernatant, leaving the cell pellet.
4. Freeze at -20°C and transfer at -80°C for long-term storage.

#### Results

After cell isolation from blood samples, cellular yields for each isolated cell type (EDTA PBMCs, Dextran PBMCs and PMNs) were calculated. The total number of cells isolated was divided by the quantity of blood used for isolation to get comparable concentrations between groups. PBMCs were isolated in two different ways, the first using the plasma-free blood collected in EDTA tubes, and the second following dextran-sedimentation of erythrocytes also from EDTA blood. PBMCs were at a higher concentration in BD groups compared to QC (**Fig. S6A-B**) for both HIV- and HIV+ subjects for EDTA PBMCs, and only HIV- subjects for Dextran PBMCs, while PMNs were higher in QC compared to BD (**Fig. S6C**). All these cells are stored at -150°C for future use.

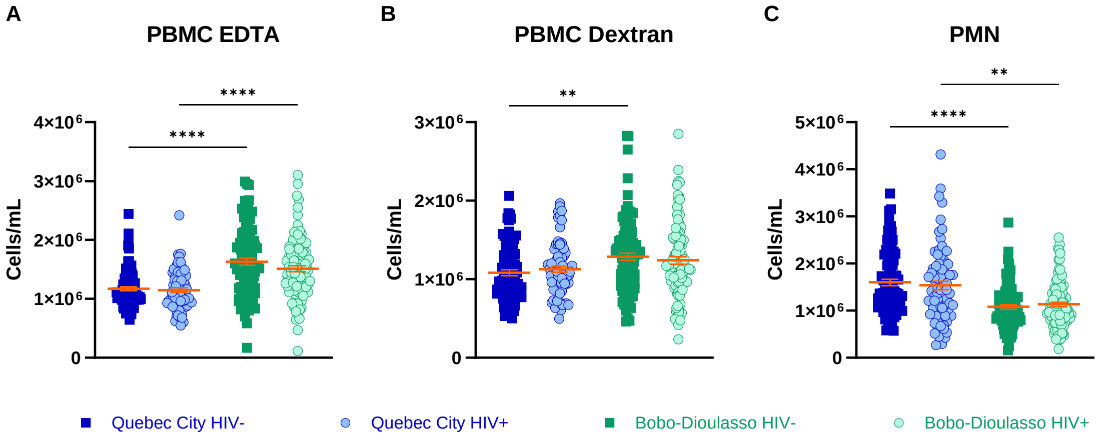


**Supplementary Figure 6. Cellular yields from blood cell isolation protocol. (A)** Yields for EDTA PBMC. **(B)** Yields for Dextran PBMC. **(C)** Yields for PMN. Quebec City HIV- n=87, HIV+ n=73; Bobo-Dioulasso HIV- n=99, HIV+ n=100. Kruskal-Wallis test, Dunn's multiple comparisons test. Data are shown as mean ± SEM. ** $p < 0.01$, **** $p < 0.0001$.

In order to compare blood cell counts and cell isolation and to evaluate the method between both locations, correlations were made between PBMCs, PMN, and the main cell types they contain. We found strong correlations between EDTA PBMCs and lymphocytes, as well as PMNs and neutrophils for QC participants, and moderate correlations for BD participants (**Fig. S7A, C**). Dextran PBMCs also correlated moderately with lymphocytes for both QC and BD (**Fig. S7D**). There were no correlations between EDTA PBMCs nor Dextran PBMCs with monocytes (**Fig. S7B, E**).

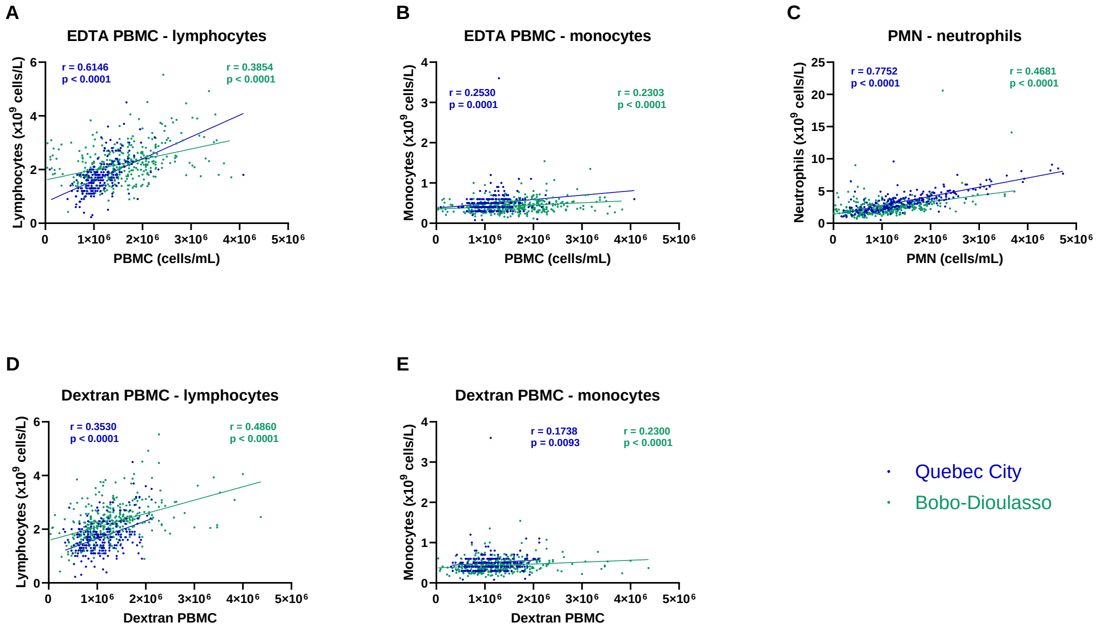


**Supplementary Figure 7. Correlations between blood cell counts and isolated cells from collected blood. (A)** EDTA PBMC and lymphocytes. QC n=223, BD n=379. **(B)** EDTA PBMC and monocytes. QC n=223, BD n=379. **(C)** PMN and neutrophils. QC n=227, BD n=380. **(D)** Dextran PBMC and lymphocytes. QC n=223, BD n=380. **(E)** Dextran PBMC and monocytes. QC n=223, BD n=380. Two-tailed Spearman correlation with 95% confidence interval was used.
